# Carbapenem-resistant Organisms in Companion Animals in New York City, 2019–2022

**DOI:** 10.1101/2025.03.16.25324050

**Authors:** Caroline A. Habrun, William G. Greendyke, Donald Szlosek, Andy Plum, Molly M. Kratz, Elise Mantell, Karen A. Alroy

## Abstract

The U.S. Centers for Disease Control and Prevention warns that carbapenem-resistant organisms (CRO) threaten human health. CRO can infect or colonize dogs and cats, with potential for zoonotic transmission to humans, but CRO prevalence in pet populations is not well-characterized. To characterize CRO prevalence among gram-negative cultured isolates from New York City (NYC) dogs and cats, we analyzed antimicrobial susceptibility data from one commercial veterinary diagnostic laboratory serving NYC veterinarians during 2019–2022. Among 16,115 gram-negative isolates, 256 (1.6%) were CRO cultured from 241 dogs and cats. CRO detections and percent positivity fluctuated during 2019–2022, including some spatial patterning. While CRO detections among companion animals were rare, increases in carbapenem-resistant *Klebsiella* spp. and *Enterobacter* spp. isolates may represent local clusters. Public health partnerships with commercial veterinary diagnostic laboratories create opportunities to share data to improve veterinary outreach and control of CRO in companion animals.

## Introduction

The emergence and spread of antimicrobial-resistant bacteria are pressing public health challenges affecting both human and animal health. Antimicrobials are essential for treating infectious diseases, and when antibiotics are no longer effective, infections can become difficult to treat. Antimicrobial-resistant bacteria were associated with approximately 4.95 million human deaths globally in 2019 [1]. While many animal species can be infected or colonized by resistant bacteria, much remains unknown about the global impact of resistant bacteria on animal health [2–5]. Since antimicrobial resistance (AR) is a multi-faceted challenge, collaborations and data sharing between human and animal health sectors are necessary to understand and control AR in humans and animals [6].

Carbapenem-resistant organisms (CRO) are an emerging type of resistant bacteria that are concerning to public health. CRO include gram-negative bacteria resistant to carbapenem antibiotics, such as meropenem, imipenem, and ertapenem. Due to their broad spectrum of activity and efficacy against many pathogens, carbapenems are primarily used to treat severe bacterial infections in people [7]. CRO infections in humans are often associated with receiving care in acute and long-term healthcare settings and can lead to severe illness, treatment failures, and even death [8]. Carbapenems are not approved for use in veterinary medicine, although they are used occasionally in companion animals in an extra-label fashion to treat multidrug-resistant infections [9]. The World Health Organization (WHO) lists carbapenems as critically important for human medicine and highlights that the sustained efficacy of these drugs is essential for humankind [8].

While there are numerous bacteria within the category of CRO, the detection of certain organisms or resistance mechanisms helps to guide the public health response to CRO. The Centers for Disease Control and Prevention (CDC) ranks different resistant bacteria according to their public health threat level: carbapenem-resistant Enterobacterales (CRE), including *Escherichia coli* and *Klebsiella pneumoniae*, and carbapenem-resistant *Acinetobacter baumannii* (CRAB) are urgent public health threats, and multidrug-resistant *Pseudomonas aeruginosa* is considered a serious threat to public health [7]. Several mechanisms result in carbapenem resistance [10]. Carbapenemases, a type of enzyme that inactivates carbapenems, comprise the most concerning resistance mechanism because genes encoding carbapenemases are frequently carried on mobile genetic elements (e.g., plasmids), facilitating rapid spread within and between bacterial populations [11].

While CRO have been detected in companion animals, public health entities are seldom notified about these detections and therefore are unable to pursue phenotypic testing for carbapenemase production or molecular identification of carbapenemase genes [12]. CDC has developed state and local health department guidance in humans on how to prevent and contain novel and targeted multidrug resistant organisms (MDRO), including CRO, and prioritization of public health response efforts relies on knowing the phenotypic and molecular characteristics related to the resistance mechanisms [13, 14]. Without notification and additional tests for CRO, public health entities are limited in their ability to support veterinary professionals with CRO infection prevention and control efforts.

The New York City (NYC) Health Department oversees the health of a large urban jurisdiction known for being an epicenter for emerging resistant organisms [15–16]. To establish a baseline understanding of CRO among companion animals in our jurisdiction, the NYC Health Department collaborated with a large commercial veterinary diagnostic laboratory to retrospectively analyze companion animal (i.e., dog and cat) antimicrobial susceptibility testing (AST) data for specimens submitted by NYC veterinarians during a four-year period. Partnering with a commercial laboratory offered a high volume of specimens submitted for testing, a laboratory catchment area representing animal care facilities in over 50% of NYC Zip Codes, and consistent laboratory methods used for AST. While a variety of MDRO can infect or colonize companion animals, we focused on CRO because the CDC and WHO have deemed them priority resistant organisms and because companion animals live in close proximity to their human guardians and care givers, thus enabling ample opportunity for zoonotic transmission [7, 8, 17]. We aimed to illustrate the need for systematic animal CRO surveillance in NYC and ultimately improve infection prevention and control for humans and their animal companions.

## Methods

We evaluated all bacterial isolates tested at a single commercial veterinary diagnostic laboratory for culture and antimicrobial susceptibility from dog and cat specimens submitted by veterinarians in NYC during January 1, 2019–December 31, 2022. Specimens were submitted from veterinary clinics, animal care shelters, and veterinary referral hospitals. Each isolate was associated with a unique deidentified patient number, patient species, patient age, year of sample submission, five-digit ZIP Code of submitting facility, the source where the sample was obtained from the patient (e.g., general specimen source information such as nasal, skin, etc.), the site where the sample was taken from the patient (e.g., more specific source information, such as nasal flush, bite wound, etc.), organism identified, and the organism’s AST result for carbapenems tested at this laboratory, imipenem and meropenem. Antimicrobial susceptibility status was reported as “susceptible”, “resistant”, or “intermediate” using clinical breakpoint values established by the Clinical Laboratory Standards Institute [18]. Susceptibility testing was performed using techniques including the Vitek® (bioMérieux) automated system [19] and Kirby-Bauer testing [20].

Data were cleaned, deduplicated, and analyzed using SAS Enterprise Guide 8.3 (SAS Institute, Cary NC). We conducted descriptive analyses and described the epidemiologic characteristics and temporal patterns of CRO in dogs and cats. Isolates where only a fungal organism was detected and isolates with an environmental source were removed from the dataset. Many patients had multiple or repeat cultures, so to avoid overestimating CRO prevalence, we deduplicated isolates. Isolates were considered a duplicate if they shared the same unique patient number and same organism by genus (Figure 1). Multiple isolates were attributed to one patient if a different genus and species were detected. Initial CRO detections, identified as isolates with an earlier collection year were retained during deduplication.

**Figure 1:**
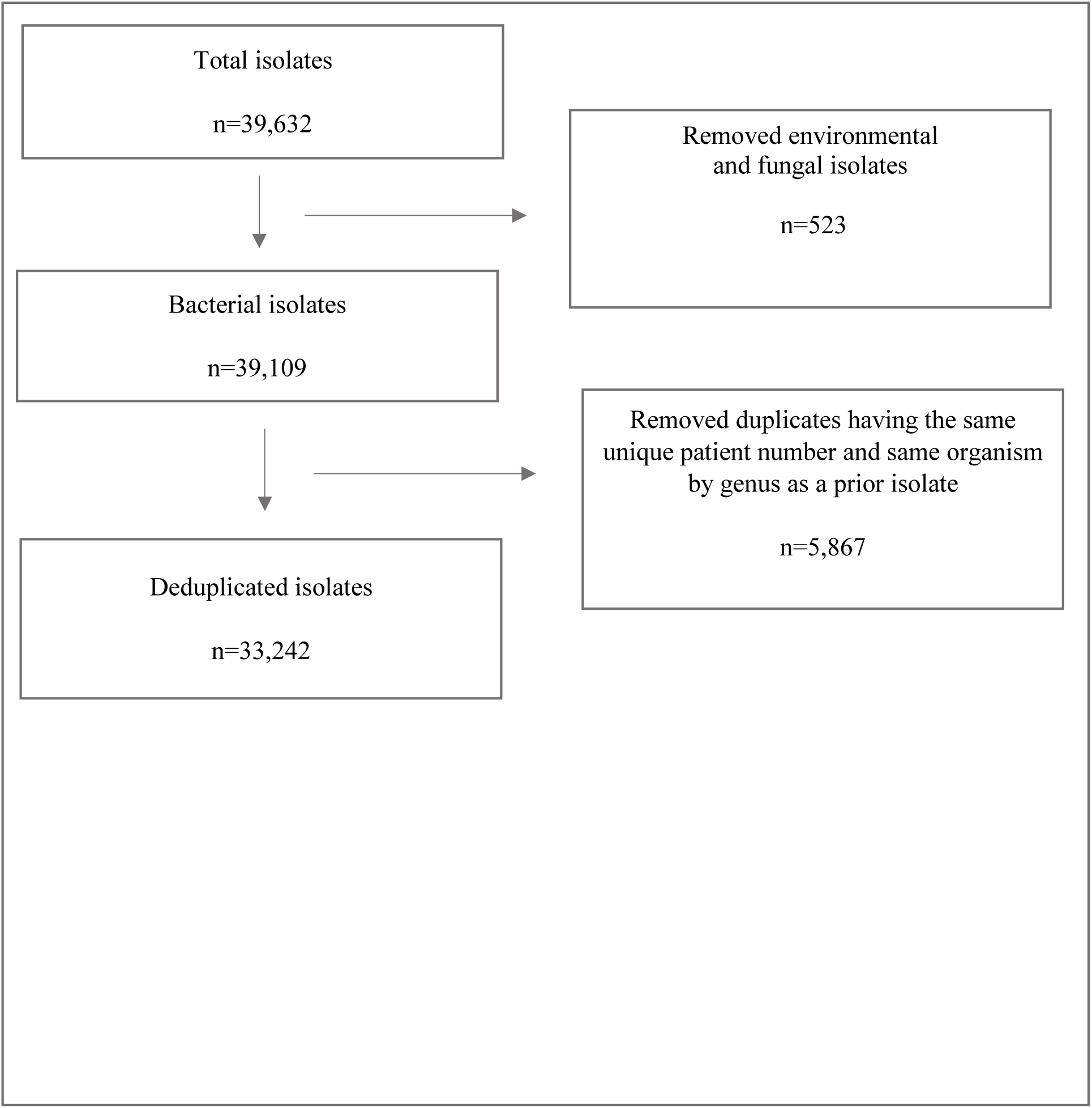
Diagram describing data de-duplication of isolates identified by culture and antimicrobial susceptibility testing by animal care facilities in New York City from a single commercial diagnostic laboratory, 2019–2022.

A CRO isolate was defined as a gram-negative bacterial isolate with AST results indicating resistance to either meropenem and/or imipenem. Since carbapenems are typically used to treat resistant gram-negative infections, and carbapenem-resistance is a clinical concern for these bacteria, we restricted the analysis to gram-negative bacteria, specifically bacterial species without intrinsic resistance to carbapenems [18]. All organisms were categorized and described by genus and species (Supplementary Table 2).

Age data were categorized in 5 groups, based on age groups established for cats: 0–4 (youth), 5–9 (early midlife), 10–11 (late midlife), 12–13 (senior) and 14–25 years (geriatric) [21]. Dogs and cats with missing ages and those aged >25 years were categorized as age unknown; the latter were likely data errors and not reflecting a true age. We used cat age groups for both species because these life stages are appropriate for cats and small and medium breed dogs (just not large breed dogs). Breed and size, which determine canine life stages, were unavailable.

The variable “composite site” combined data from both source and site. Composite site was comprised of 12 non-sterile and 9 normally sterile categories, as well as other and unknown (Supplementary Table 1) [22]. Urinary samples were not considered sterile [23, 24]. If source was “other”, the site variable was used to define composite site, if possible. The composite site category of “other” included sources that were too vague to allow for categorization, and “unknown” included samples with a missing or unknown source.

## Results

After removing environmental and fungal isolates and deduplication, 33,242 bacterial isolates remained for analysis (Figure 1). The dataset included 25,848 and 7,394 bacterial isolates cultured from dogs and cats respectively, with AST results representing 21,242 unique canine and feline patients during 2019–2022. Of 33,242 isolates during the four-year period, 16,115 (49%) represented gram-negative bacteria.

Of the 16,115 gram-negative isolates, 256 (1.6%) were resistant to imipenem, meropenem, or both. These 256 CRO represented specimens collected from 241 individual animals, specifically, 180 dogs and 61 cats. There were no differences in the overall prevalence between cats and dogs. Animals with CRO ranged in age from 0–17 years, with a mean of 7.3 years in dogs and 9.7 years in cats. Bacterial isolates cultured from early midlife animals (5–9 years) represented 37.4% of canine and 31.8% of feline CRO (Table 1).

**Table 1:**
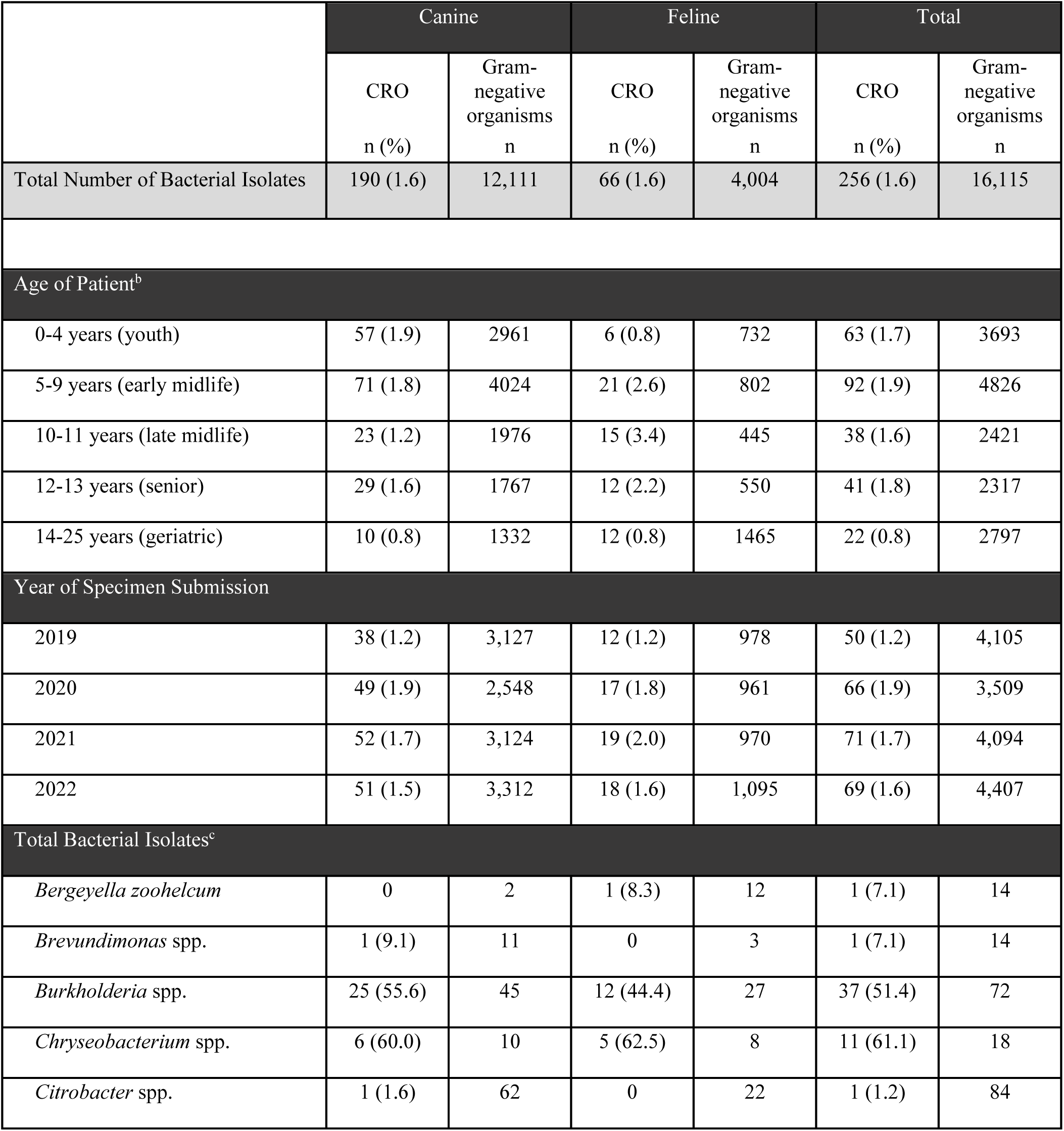
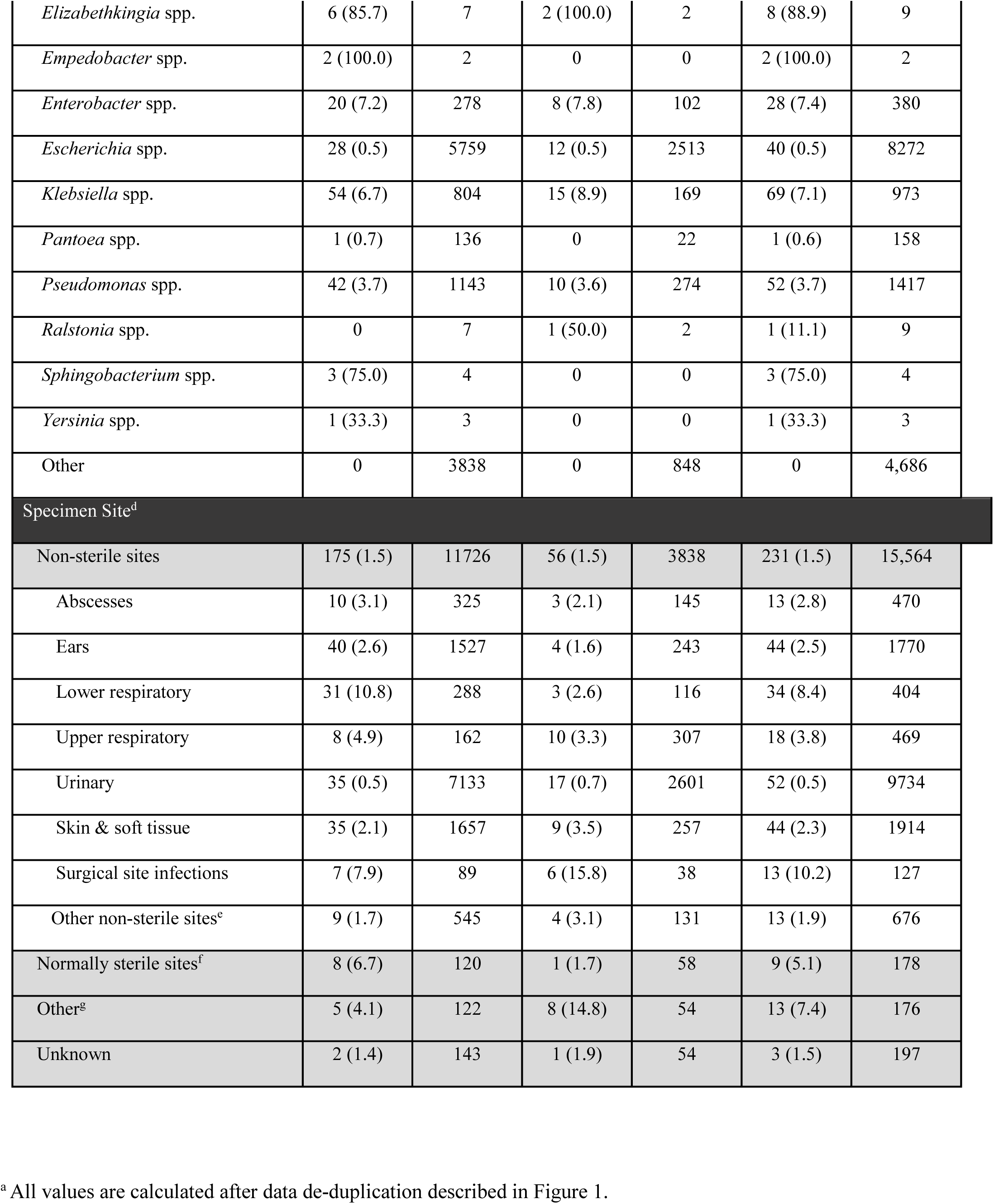
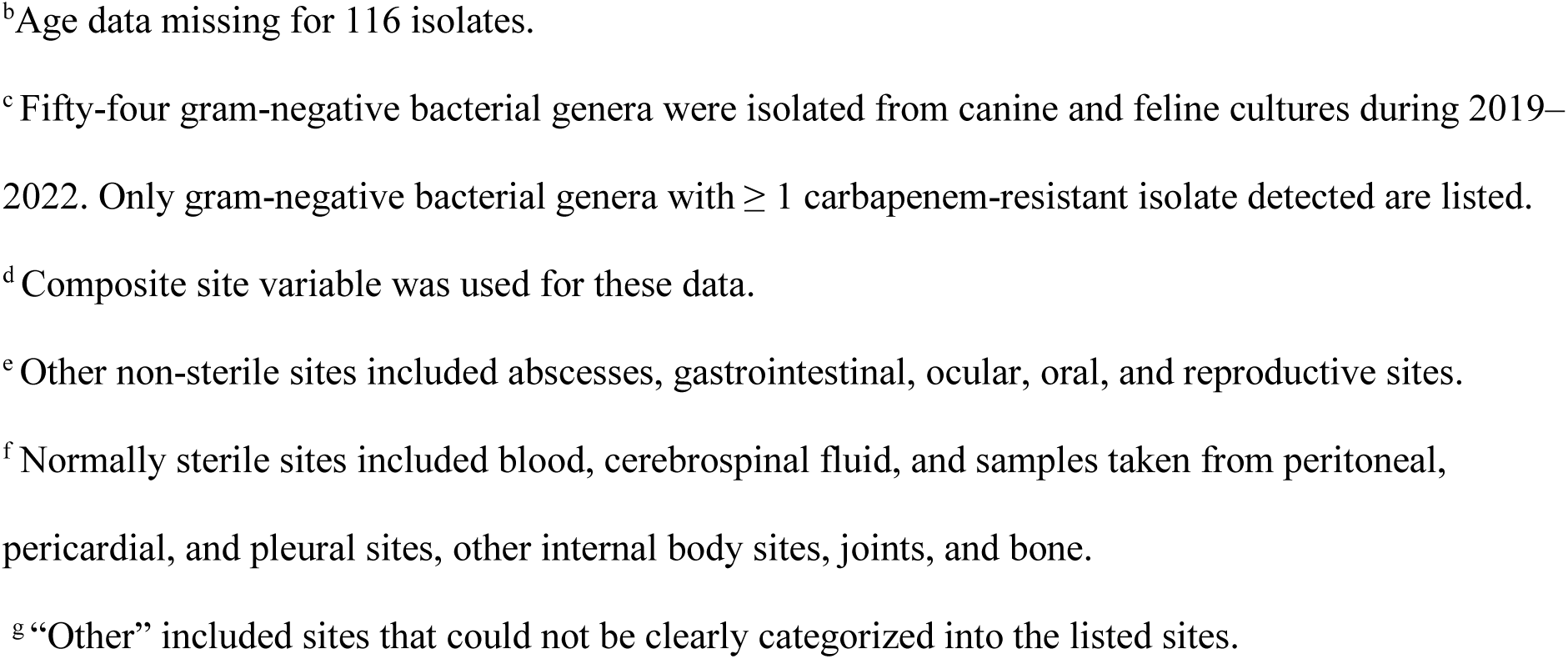
Distribution of isolates of gram-negative and carbapenem-resistant organisms (CRO)^a^ identified by culture and antimicrobial susceptibility testing by animal care facilities in New York City from a single commercial diagnostic laboratory, 2019–2022.

|  | Canine |  | Feline |  | Total |  |
| --- | --- | --- | --- | --- | --- | --- |
|  | CRO<br>n (%) | Gram-negative<br>organisms<br>n | CRO<br>n (%) | Gram-negative<br>organisms<br>n | CRO<br>n (%) | Gram-negative<br>organisms<br>n |
| Total Number of Bacterial Isolates | 190 (1.6) | 12,111 | 66 (1.6) | 4,004 | 256 (1.6) | 16,115 |
| Age of Patient<sup>b</sup> |  |  |  |  |  |  |
| 0-4 years (youth) | 57 (1.9) | 2961 | 6 (0.8) | 732 | 63 (1.7) | 3693 |
| 5-9 years (early midlife) | 71 (1.8) | 4024 | 21 (2.6) | 802 | 92 (1.9) | 4826 |
| 10-11 years (late midlife) | 23 (1.2) | 1976 | 15 (3.4) | 445 | 38 (1.6) | 2421 |
| 12-13 years (senior) | 29 (1.6) | 1767 | 12 (2.2) | 550 | 41 (1.8) | 2317 |
| 14-25 years (geriatric) | 10 (0.8) | 1332 | 12 (0.8) | 1465 | 22 (0.8) | 2797 |
| Year of Specimen Submission |  |  |  |  |  |  |
| 2019 | 38 (1.2) | 3,127 | 12 (1.2) | 978 | 50 (1.2) | 4,105 |
| 2020 | 49 (1.9) | 2,548 | 17 (1.8) | 961 | 66 (1.9) | 3,509 |
| 2021 | 52 (1.7) | 3,124 | 19 (2.0) | 970 | 71 (1.7) | 4,094 |
| 2022 | 51 (1.5) | 3,312 | 18 (1.6) | 1,095 | 69 (1.6) | 4,407 |
| Total Bacterial Isolates<sup>c</sup> |  |  |  |  |  |  |
| *Bergeyella zoohelcum* | 0 | 2 | 1 (8.3) | 12 | 1 (7.1) | 14 |
| *Brevundimonas* spp. | 1 (9.1) | 11 | 0 | 3 | 1 (7.1) | 14 |
| *Burkholderia* spp. | 25 (55.6) | 45 | 12 (44.4) | 27 | 37 (51.4) | 72 |
| *Chryseobacterium* spp. | 6 (60.0) | 10 | 5 (62.5) | 8 | 11 (61.1) | 18 |
| *Citrobacter* spp. | 1 (1.6) | 62 | 0 | 22 | 1 (1.2) | 84 |
| *Elizabethkingia* spp. | 6 (85.7) | 7 | 2 (100.0) | 2 | 8 (88.9) | 9 |
| *Empedobacter* spp. | 2 (100.0) | 2 | 0 | 0 | 2 (100.0) | 2 |
| *Enterobacter* spp. | 20 (7.2) | 278 | 8 (7.8) | 102 | 28 (7.4) | 380 |
| *Escherichia* spp. | 28 (0.5) | 5759 | 12 (0.5) | 2513 | 40 (0.5) | 8272 |
| *Klebsiella* spp. | 54 (6.7) | 804 | 15 (8.9) | 169 | 69 (7.1) | 973 |
| *Pantoea* spp. | 1 (0.7) | 136 | 0 | 22 | 1 (0.6) | 158 |
| *Pseudomonas* spp. | 42 (3.7) | 1143 | 10 (3.6) | 274 | 52 (3.7) | 1417 |
| *Ralstonia* spp. | 0 | 7 | 1 (50.0) | 2 | 1 (11.1) | 9 |
| *Sphingobacterium* spp. | 3 (75.0) | 4 | 0 | 0 | 3 (75.0) | 4 |
| *Yersinia* spp. | 1 (33.3) | 3 | 0 | 0 | 1 (33.3) | 3 |
| Other | 0 | 3838 | 0 | 848 | 0 | 4,686 |
| Specimen Site<sup>d</sup> |  |  |  |  |  |  |
| Non-sterile sites | 175 (1.5) | 11726 | 56 (1.5) | 3838 | 231 (1.5) | 15,564 |
| Abscesses | 10 (3.1) | 325 | 3 (2.1) | 145 | 13 (2.8) | 470 |
| Ears | 40 (2.6) | 1527 | 4 (1.6) | 243 | 44 (2.5) | 1770 |
| Lower respiratory | 31 (10.8) | 288 | 3 (2.6) | 116 | 34 (8.4) | 404 |
| Upper respiratory | 8 (4.9) | 162 | 10 (3.3) | 307 | 18 (3.8) | 469 |
| Urinary | 35 (0.5) | 7133 | 17 (0.7) | 2601 | 52 (0.5) | 9734 |
| Skin & soft tissue | 35 (2.1) | 1657 | 9 (3.5) | 257 | 44 (2.3) | 1914 |
| Surgical site infections | 7 (7.9) | 89 | 6 (15.8) | 38 | 13 (10.2) | 127 |
| Other non-sterile sites<sup>e</sup> | 9 (1.7) | 545 | 4 (3.1) | 131 | 13 (1.9) | 676 |
| Normally sterile sites<sup>f</sup> | 8 (6.7) | 120 | 1 (1.7) | 58 | 9 (5.1) | 178 |
| Other<sup>g</sup> | 5 (4.1) | 122 | 8 (14.8) | 54 | 13 (7.4) | 176 |
| Unknown | 2 (1.4) | 143 | 1 (1.9) | 54 | 3 (1.5) | 197 |

<sup>a</sup> All values are calculated after data de-duplication described in Figure 1.
<sup>b</sup>Age data missing for 116 isolates.
<sup>c</sup>Fifty-four gram-negative bacterial genera were isolated from canine and feline cultures during 2019–2022. Only gram-negative bacterial genera with $\geq 1$ carbapenem-resistant isolate detected are listed.
<sup>d</sup>Composite site variable was used for these data.
<sup>e</sup>Other non-sterile sites included abscesses, gastrointestinal, ocular, oral, and reproductive sites.
<sup>f</sup>Normally sterile sites included blood, cerebrospinal fluid, and samples taken from peritoneal, pericardial, and pleural sites, other internal body sites, joints, and bone.
<sup>g</sup>“Other” included sites that could not be clearly categorized into the listed sites.

CRO detections increased from 2019 to 2021, with a 32% increase in 2020 from the previous year. Percent positivity of carbapenem-resistant isolates increased from 1.2% in 2019 to 1.9% in 2020, then declined to 1.6% in 2022. (Figure 2). The most frequently cultured CRO were *Klebsiella* spp., *Pseudomonas aeruginosa*, and *Escherichia coli* (Table 1). CRO isolates from four bacterial genera, *Klebsiella* spp., *Pseudomonas* spp., *Elizabethkingia* spp. and *Enterobacter* spp. demonstrated an overall increase in isolates over time, while resistant isolates from all other genera had an overall decrease (Figure 3). Some bacterial species showed carbapenem resistance in >50% of isolates, including *Empedobacter* spp. (2/2, 100%), *Elizabethkingia* spp. (8/9, 89%), *Sphingobacterium* spp. (3/4, 75%), *Chryseobacterium* spp. (11/18, 61%), and *Burkholderia* spp. (37/72, 51%).

**Figure 2:**
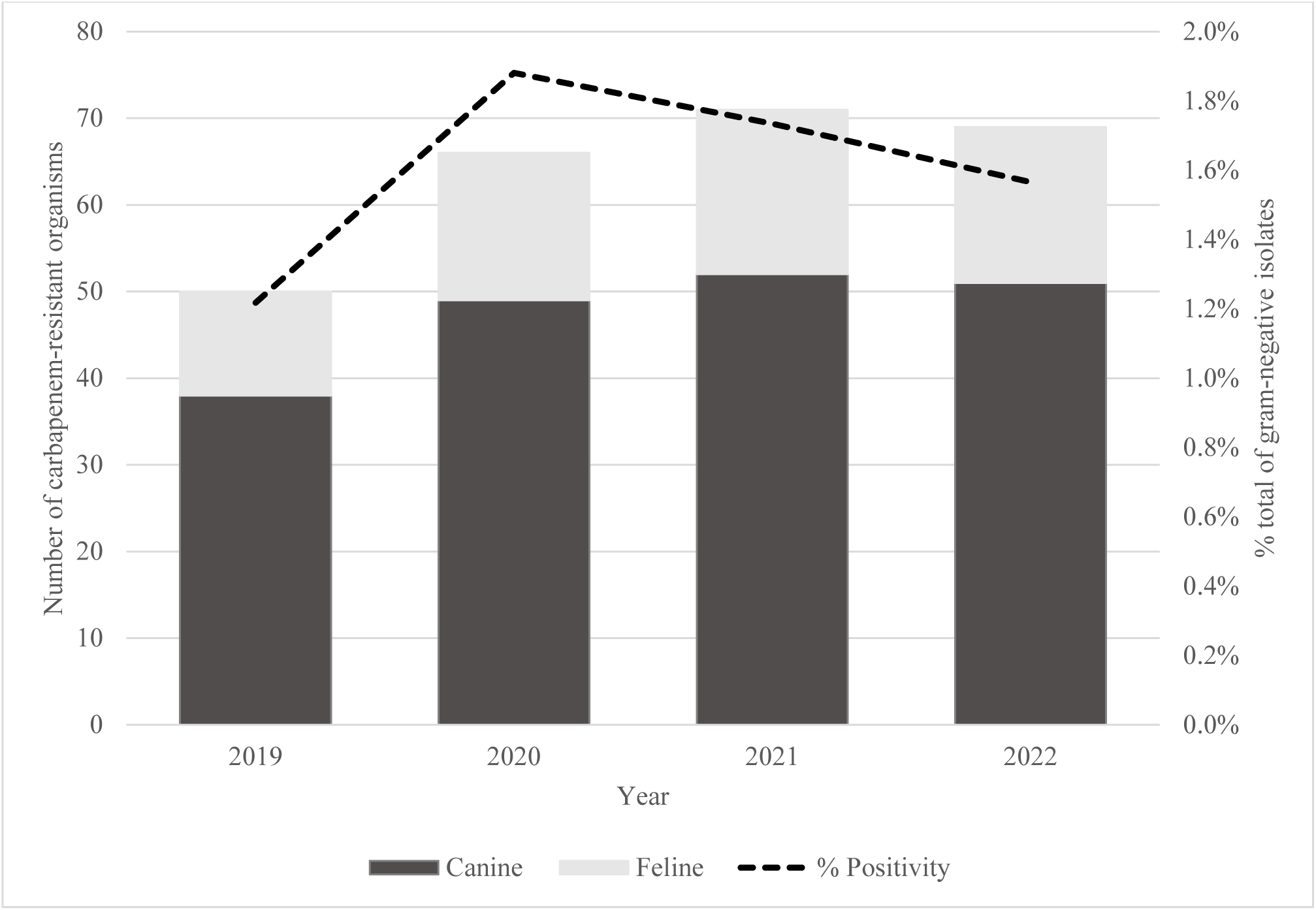
Number of isolates of carbapenem-resistant organisms (CRO) identified by culture and antimicrobial susceptibility testing by animal care facilities in New York City from a single commercial diagnostic laboratory, 2019–2022, and percent of gram-negative isolates testing positive as a CRO. Bars indicate total numbers of isolates of CRO by species and year. Dotted line indicates carbapenem-resistance percent positivity by year.

**Figure 3:**
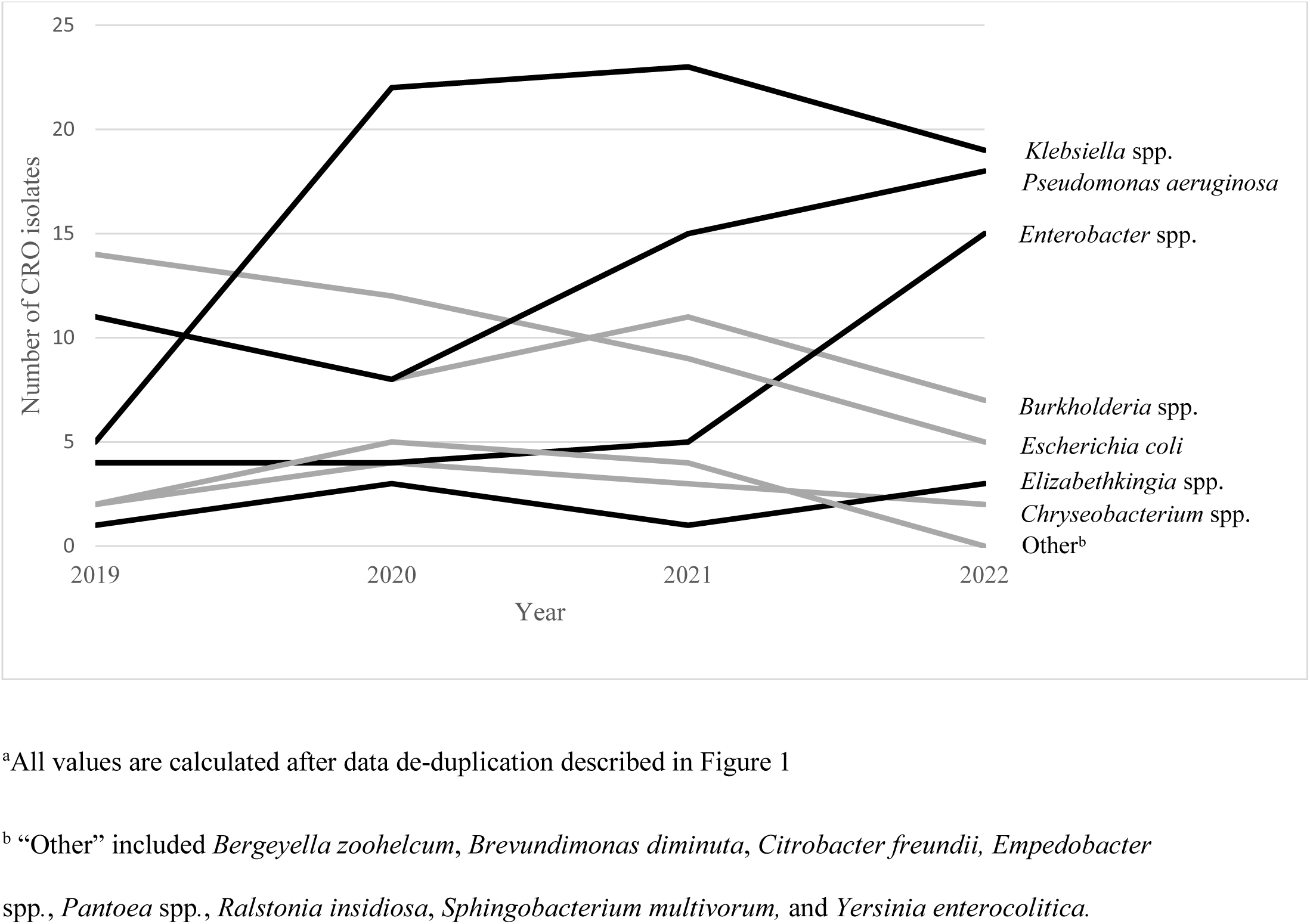
Number of isolates of carbapenem-resistant organisms (CRO) identified by culture and antimicrobial susceptibility testing by animal care facilities in New York City from a single commercial diagnostic laboratory, 2019–2022. Black lines represent pathogens demonstrating an overall increase in isolates over time. Gray lines represent pathogens demonstrating an overall decrease in isolates over time^a^.

Most cultures from companion animals were sampled from non-sterile specimen sites (15,564/16,115, 96.6%). However, of the 256 CRO, there was a higher proportion of CRO detected in normally sterile sites (9/178, 5.1%) compared with non-sterile sites (231/15,564, 1.5%) (Table 1). The most common specimen sites where CRO were detected were urinary (52/256, 20.3%), skin and soft tissue (44/256, 17.1%), ears (44/256, 17.2%), and lower respiratory (35/256, 13.7%) (Table 1). Carbapenem-resistant *Pseudomonas aeruginosa* and *Burkholderia* spp. were most often cultured from ears, and carbapenem-resistant *Klebsiella* spp. and *E. coli* were most often cultured from urinary sources (Supplementary Table 3). Most of the CRO cultured from sterile sites (8/9, 88.9%) were cultured from dogs. Only *Escherichia coli, Enterobacter* spp., and *Klebsiella* spp. were cultured from normally sterile sites (Figure 4), and these were cultured from peritoneal fluid (5/9, 55.6%), internal body sites (2/9, 22.2%), and blood (1/9, 11.1%). The remaining isolates were either cultured from other sites (5.1%) or unknown (1.2%).

**Figure 4:**
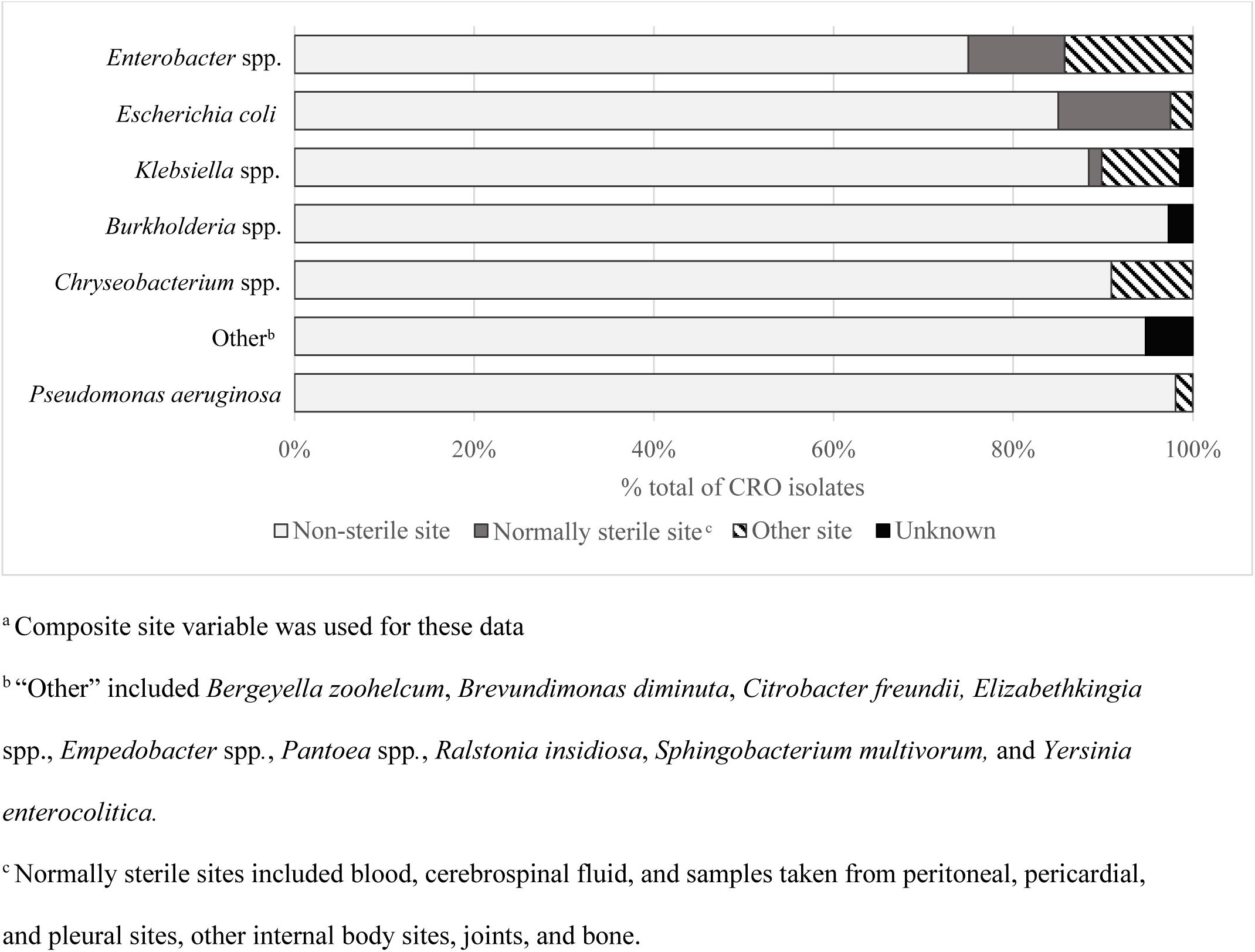
Percentage of carbapenem-resistant organisms (CRO) isolates identified by culture and antimicrobial susceptibility testing by animal care facilities in New York City from a single commercial diagnostic laboratory, by non-sterile or normally sterile specimen site^a^ and genus, 2019–2022.

Geographic patterns were observed in the data. Veterinarians from a single ZIP Code submitted 21% (3,369/16,115) of bacterial cultures that grew gram-negative isolates. Among all cultures submitted citywide, nearly 90% (62/69) of carbapenem-resistant *Klebsiella* spp. isolates and 85.7% (24/28) of carbapenem-resistant *Enterobacter* spp. isolates were submitted from within this single ZIP Code. The increase from 50 to 66 isolates (32% increase) from 2019 to 2020 was driven by a rise and sustained number of *Klebsiella* spp. isolates among primarily dogs receiving care in this ZIP Code. In addition, 93.3% (14/15) of the carbapenem-resistant *Enterobacter* spp. isolates that resulted in the sharp increase of *Enterobacter* spp. from 2021 to 2022 (Figure 3) were also submitted from this same ZIP Code. Geographic patterning was identified within two other ZIP Codes. Veterinarians from these ZIP Codes submitted 2.4% (379/16,115) and 1.3% (201/16,115) of bacterial cultures that grew gram-negative isolates, respectively. Over 45% of *Burkholderia* spp. and *Elizabethkingia* spp. CRO were submitted by one or multiple animal care facilities within each ZIP Code. Conversely, the rise in *Pseudomonas aeruginosa* isolates from 2020 to 2022 did not show any geographic patterns.

## Discussion

Of gram-negative isolates from companion animal specimens submitted by NYC veterinarians, 1.6% were CRO; this prevalence is consistent with other studies [3, 25]. Carbapenem-resistant *Klebsiella* spp., *Pseudomonas* spp., and *Enterobacter* spp. increased during 2019–2022. The upward trend of *Klebsiella* spp. and *Enterobacter* spp. detected from a single ZIP Code was likely from one or multiple veterinary referral facilities within this ZIP Code, since veterinary referral facilities are more likely to submit diagnostic bacterial cultures than general practice clinics, referral facilities treat medically complex cases with animals who have often failed first-line treatment, and referral facility patients are more likely than the general animal population to be infected or colonized with CRO [26–29]. The two other notable ZIP Codes with high-proportions of carbapenem-resistant *Burkholderia* spp. and *Elizabethkingia* spp. could also represent bacterial cultures submitted from referral facilities. Without a timely animal CRO surveillance system in NYC that could enable the provision of additional epidemiologic data or molecular characterization of the isolates, we cannot exclude the possibility of a local cluster or within-facility transmission. The increase in carbapenem-resistant *Pseudomonas aeruginosa* without a geographic focus could potentially be due to contaminated products [30]; however, similarly, this could not be determined without additional information. Above all, these results highlight that sharing data via partnerships between public health and commercial veterinary diagnostic laboratories can inform surveillance and ultimately might help limit the spread of CRO in animals and people.

Information on companion animal CRO has largely been gathered from outbreak investigations [26, 27, 30, 31]. To date, there have been four notable CRO companion animal U.S. outbreaks, including clusters at two veterinary referral hospitals [26, 27], one at an animal rescue facility [31], and a cluster associated with contaminated products [30]. Most clusters were detected through a laboratory associated with the Food and Drug Administration (FDA)’s Laboratory Investigation and Response Network (Vet-LIRN). FDA’s Vet-LIRN supports state and academic veterinary laboratories to track AR in bacteria from sick animals; however, not all animals with a CRO become visibly sick. CRO colonization, when animals carry CRO in or on their bodies without showing signs of illness, can lead to environmental spread and direct transmission to other pets and people [32]. Spread via contaminated equipment in veterinary hospitals [27] and in community areas such as city parks [33] has been documented.

In addition to outbreak investigations, laboratory-based studies have also characterized CRO in animals. Vet-LIRN laboratories in Kansas and Missouri had sporadic CRO companion animal detections over several years [3], and a nationwide study using commercial veterinary diagnostic data for Enterobacterales found that 0.76% of isolates were intermediate and 0.38% were resistant to imipenem [25].

Many animals with a CRO in our dataset were in the early midlife age category (aged 5–9 years), in contrast to CRO detections in humans which most often occur later in life. A recent study of CRE across the United States found that human patients’ median age was 69 years, and most patients were aged 65–79 years [34]. Age differences between human and animal populations could reflect differences in how humans versus animals are exposed to CRO. For example, there is no animal equivalent to long-term care facilities, which are associated with CRO exposure in humans [7, 8].

Common CRO in companion animals cultured from non-sterile and sterile sites are associated with different types of infections, with various causes. The most common CRO isolated from companion animals, *Klebsiella* spp*., E. coli,* and *Pseudomonas aeruginosa*, were most often isolated from non-sterile sites, which could indicate infections by normal flora or contamination of a damaged body site, such as a skin abrasion [35]. These bacteria can be the cause of urinary tract infections, pyoderma, and otitis externa [36, 37], and the use of empiric antibiotic treatment for these conditions is common [38–39]. Recurrent antibiotic treatment without AST could potentially lead to treatment failures, adverse side effects, and selection of resistant bacteria. In contrast, the most common organisms cultured from normally sterile sites included *E. coli, Enterobacter* spp., and *Klebsiella* spp., and peritoneal fluid was the most common sterile site for CRO isolation. Peritonitis in companion animals may result from various causes, such as hematogenous spread or damage to the gastrointestinal or urogenital tract [40].

Some less commonly cultured genera showed a high proportion of carbapenem resistance, including *Empedobacter* spp., *Elizabethkingia* spp., *Sphingobacterium* spp., *Chryseobacterium* spp., and *Burkholderia* spp. The genera *Elizabethkingia* spp. and *Chryseobacterium* spp. have been recently described as emerging infections in human hospitals and long-term care settings [41, 42], whereas *Burkholderia* spp. are water-related opportunistic organisms that are historically known to be associated with environmental contamination within healthcare settings and nosocomial infections [43, 44]. In companion animal veterinary settings, *Burkholderia* spp. has been found in contaminated chlorhexidine 2% scrub and associated with nosocomial infections in cats [45]. Given the sporadic detections of these genera, high proportion of carbapenem resistance observed, and their potential for nosocomial spread, veterinarians and public health partners should carefully review AST data and consider the potential for within-hospital transmission when these organisms are cultured.

Our findings are subject to at least five limitations. First, we assessed CRO among gram-negative isolates cultured from specimens submitted by veterinarians. This likely overestimated CRO in the general population of companion animals residing in NYC, as animals with recurrent or chronic conditions are more likely to have AST performed [38–39]. Animals colonized with CRO or with mild or self-limiting infections may not be detected or offered AST. Our deduplication process did not account for animals infected by two distinct CRO strains of the genus and species which could only be distinguished with genetic sequencing. Although our methodology overall is more likely to have overestimated CRO prevalence, rather than underestimated. Second, our study period overlapped with the emergency phase of the COVID- 19 pandemic. Although there were anecdotal reports of increases in demand for veterinary services during the COVID-19 pandemic [46], there is limited available literature to know if this was widespread, sustained, or if this fluctuated and how this may have impacted CRO detections during 2020–2022. Third, high veterinary costs may further limit testing since bacterial cultures are typically paid for by pet owners, potentially over-representing pets living in households with a higher socioeconomic status. Lower socioeconomic communities have disproportionately been impacted by resistant bacteria, yet pets from households unable to pay for bacterial cultures were not included in our dataset [47]. Fourth, our data source was a single commercial veterinary diagnostic laboratory and thus does not represent sampling at all animal care facilities in NYC. While we cannot quantify the number of facilities that utilize this laboratory, it is considered one of two veterinary laboratories that dominate the industry [48]. Fifth, our dataset was at the geographic resolution of ZIP Code of submitting facility. Thus, we were unable to distinguish whether prevalence increases within a ZIP Code were attributable to one or multiple animal care facilities.

Companion animal AR research has focused primarily on gram-positive bacteria [17], although research on resistant gram-negative bacteria, and CRO in particular, is growing [27, 30, 31, 49]. CRO data provided at broader geographic scales, either a diagnostic laboratory catchment population [3], or a national or municipal jurisdiction [25], can potentially be used to identify CRO clusters or hotspots [25]. In animal populations, delayed detection of emerging and zoonotic pathogens, and limited notification to public health entities, can ultimately result in increased transmission between animals or from animals to humans [50]. While this report and other companion animal CRO studies are retrospective [3, 25], ideally, for public health entities to prioritize and implement interventions to control CRO in veterinary settings, systematic animal CRO surveillance systems are needed to provide real-time CRO notifications, resistance mechanism testing, and facility-level data. Strengthening CRO surveillance in animals, especially in collaboration with commercial veterinary diagnostic laboratories, could improve understanding and enable public health action to help control the spread of companion animal CRO.

## Data Availability

If interested in accessing data or other materials related to this manuscript, readers may contact the corresponding author.

## Acknowledgments

Shama Desai Ahuja, Tristan D. McPherson, Sharon Greene, New York City Department of Health and Mental Hygiene.

## Disclaimer

The findings and conclusions of this paper are those of the authors and do not necessarily represent the official position of the New York City Department of Health and Mental Hygiene.

## Authorship Contribution

KAA and CAH conceptualized and designed the project. DS curated the data. CAH and EM conducted the data analysis. CAH and KAA drafted the manuscript. All authors provided intellectual contribution and reviewed and approved the final draft of the manuscript.

## References

1. Murray CJ. Global burden of bacterial antimicrobial resistance in 2019: a systematic analysis. Lancet. 2022; 399:629–55.

2. Sobkowich KE, Weese JS, Poljak Z, et al. Epidemiology of companion animal AMR in the United States of America: filling a gap in the one health approach. Front Public Health. 2023; 11:1161950.

3. KuKanich K, Burklund A, McGaughey R, et al. One health approach for reporting veterinary carbapenem-resistant Enterobacterales and other bacteria of public health concern. Emerg Infect Dis. 2023 ; 29:1–9.

4. Nobrega DB, Tang KL, Caffrey NP, et al. Prevalence of antimicrobial resistance genes and its association with restricted antimicrobial use in food-producing animals: a systematic review and meta-analysis. J Antimicrob Chemother. 2021; 76:561–75.

5. Wang J, Ma ZB, Zeng ZL, et al. The role of wildlife (wild birds) in the global transmission of antimicrobial resistance genes. Zool Res. 2017 ; 38:55–80.

6. Laborda P, Sanz-García F, Ochoa-Sánchez LE, et al. Wildlife and antibiotic resistance. Front Cell Infect Microbiol. 2022 ;12:873989.

7. Centers for Disease Control and Prevention. Antibiotic resistance threats in the United States, 2019. Available at: https://www.cdc.gov/antimicrobial-resistance/media/pdfs/2019-ar-threats-report-508.pdf.Accessed 15 April 2024.

8. World Health Organization. Critically important antimicrobials for human medicine. Available at: https://www.who.int/groups/advisory-group-on-the-who-list-of-critically-important-antimicrobials. Accessed 15 April 2024.

9. Smith A, Wayne AS, Fellman CL, et al. Usage patterns of carbapenem antimicrobials in dogs and cats at a veterinary tertiary care hospital. J Vet Intern Med. 2019 ;33:1677–85.

10. Aurilio C, Sansone P, Barbarisi M, et al. Mechanisms of action of carbapenem resistance. Antibiotics (Basel). 2022 ; 11:421.

11. Logan LK, Weinstein RA. The epidemiology of carbapenem-resistant Enterobacteriaceae: The impact and evolution of a global menace. J Infect Dis. 2017 ; 215(suppl_1):S28–s36.

12. Waltenburg MA, Shugart A, Loy JD, et al. A survey of current activities and technologies used to detect carbapenem resistance in bacteria isolated from companion animals at veterinary diagnostic laboratories-United States, 2020. J Clin Microbiol. 2022; 60:e0215421.

13. Centers for Disease Control and Prevention. Public health strategies to prevent the spread of novel and targeted multidrug-resistant organisms (MDROs). Available at: https://www.cdc.gov/healthcare-associated-infections/php/preventing-mdros/mdro-containment-strategy.html. Accessed 20 April 2024.

14. Centers for Disease Control and Prevention. Interim guidance for a public health response to contain novel or targeted multidrug-resistant organisms (MDROs). Available at: https://www.cdc.gov/healthcare-associated-infections/php/preventing-mdros/mdro-containment-strategy.html. Accessed 20 April 2024.

15. Adams E, Quinn M, Tsay S, et al. *Candida auris* in healthcare facilities, New York, USA, 2013–2017. Emerg Infect Dis. 2018; 24:1816–24.

16. Caplan AS, Chaturvedi S, Zhu Y, et al. Notes from the field: first reported U.S. cases of tinea caused by *Trichophyton indotineae* - New York City, December 2021–March 2023. MMWR Morb Mortal Wkly Rep. 2023; 72:536–7.

17. Jin, M., Osman M, Green BA, et al., Evidence for the transmission of antimicrobial resistant bacteria between humans and companion animals: A scoping review. One Health. 2023; 17:100593.

18. CLSI. Performance standards for antimicrobial disk and dilution susceptibility tests for bacteria isolated from animals. 5th ed. CLSI supplement VET01S. Clinical and Laboratory Standards Institute; 2020.

19. Pincus D. Microbial dentification using the bioMérieux VITEK2 system.: Hazelwood, MI: Encyclopedia of Rapid Microbiological Methods; 2006.

20. Biemer JJ. Antimicrobial susceptibility testing by the Kirby-Bauer disc diffusion method. Ann Clin Lab Sci (1971). 1973; 3:135–40.

21. Salt C, Saito EK, O’Flynn C, et al. Stratification of companion animal life stages from electronic medical record diagnosis data. J Gerontol A Biol Sci Med Sci. 2023; 78:579–86.

22. Schuchat A, Hilger T, Zell E, et al. Active bacterial core surveillance of the emerging infections program network. Emerg Infect Dis. 2001; 7:92–9.

23. Burton EN, Cohn LA, Reinero CN, et al. Characterization of the urinary microbiome in healthy dogs. PLoS One. 2017; 12:e0177783.

24. Melgarejo T, Oakley BB, Krumbeck JA, et al. Assessment of bacterial and fungal populations in urine from clinically healthy dogs using next-generation sequencing. J Vet Intern Med. 2021; 35:1416–26.

25. Sobkowich K, Poljak Z, Weese JS, et al. Prevalence and distribution of carbapenem-resistant Enterobacterales in companion animals: A nationwide study in the United States using commercial laboratory data. J Vet Intern Med. 2024; 38:2642–53.

26. Cole SD, Peak L, Tyson GH, et al. New Delhi metallo-β-lactamase-5-producing *Escherichia coli* in companion animals, United States. Emerg Infect Dis. 2020; 26:381–3.

27. Lavigne SH, Cole SD, Daidone C, et al. Risk factors for the acquisition of a blandm-5 carbapenem-resistant Escherichia coli in a veterinary hospital. J Am Anim Hosp Assoc. 2021; 57.

28. Gentilini F, Turba ME, Pasquali F, et al. Hospitalized pets as a source of carbapenem-resistance. Front Microbiol. 2018 ; 9:2872.

29. Dazio V, Nigg A, Schmidt JS, et al. Acquisition and carriage of multidrug-resistant organisms in dogs and cats presented to small animal practices and clinics in Switzerland. J Vet Intern Med. 2021 ; 35:970–9.

30. Price ER, McDermott D, Sherman A, et al. Canine multidrug-resistant *Pseudomonas aeruginosa* cases linked to human artificial tears–related outbreak. Emerg Infect Dis. 2024 ; 30:2689–2691.

31. Minnesota Board of Animal Health. Veterinary alert: Antimicrobial resistance (super bugs) in companion animals. March 28, 2022. Available from: https://content.govdelivery.com/accounts/MNBAH/bulletins/310d6a2. Accessed 22 April 2024.

32. Centers for Disease Control and Prevention. Carbapenem-resistant Enterobacterales and veterinarian basics. Available at: https://www.cdc.gov/cre/about/veterinarians.html. Accessed 15 April 2024.

33. Haenni M, Métayer V, Lupo A, et al. Spread of the bla(OXA-48)/IncL plasmid within and between dogs in city parks, France. Microbiol Spectr. 2022; 10:e0040322.

34. Duffy N, Li R, Czaja CA, et al. Trends in incidence of carbapenem-resistant Enterobacterales in 7 US sites, 2016─2020. Open Forum Infect Dis. 2023 ;10:ofad609.

35. Nocera FP, Ambrosio M, Fiorito F, et al. On gram-positive and gram-negative bacteria-associated canine and feline skin infections: A 4-year retrospective study of the University Veterinary Microbiology Diagnostic Laboratory of Naples, Italy. Animals (Basel). 2021; 11.

36. Hernando E, Vila A, D’Ippolito P, et al. Prevalence and characterization of urinary tract infection in owned dogs and cats from Spain. Top Companion Anim Med. 2021; 43:100512.

37. Yudhanto S, Hung CC, Maddox CW, et al. Antimicrobial resistance in bacteria isolated from canine urine samples submitted to a veterinary diagnostic laboratory, Illinois, United States. Front Vet Sci. 2022; 9:867784.

38. Bollig ER, Granick JL, Webb TL, et al. A quarterly survey of antibiotic prescribing in small animal and equine practices-Minnesota and North Dakota, 2020. Zoonoses Public Health. 2022; 69:864–74.

39. Beaudoin AL, Bollig ER, Burgess BA, et al. Prevalence of antibiotic use for dogs and cats in United States veterinary teaching hospitals, August 2020. J Vet Intern Med. 2023; 37:1864–75.

40. Sykes JE. Intra-abdominal Infections. Canine and Feline Infectious Diseases 2014; 859–70.

41. Mukerji R, Kakarala R, Smith SJ, et al. *Chryseobacterium indologenes*: an emerging infection in the USA. BMJ Case Rep. 2016.

42. Ratnamani MS, Rao R. *Elizabethkingia meningoseptica*: Emerging nosocomial pathogen in bedside hemodialysis patients. Indian J Crit Care Med. 2013;17:304–7.

43. Häfliger E, Atkinson A, Marschall J. Systematic review of healthcare-associated *Burkholderia cepacia* complex outbreaks: presentation, causes and outbreak control. Infect Prev Pract. 2020 ; 2:100082.

44. Vazquez Deida AA, Spicer KB, McNamara KX, et al. *Burkholderia multivorans* infections associated with use of ice and water from ice machines for patient care activities - four hospitals, California and Colorado, 2020–2024. MMWR Morb Mortal Wkly Rep. 2024 ;73:883–7.

45. Wong JK, Chambers LC, Elsmo EJ, et al. Cellulitis caused by the *Burkholderia cepacia* complex associated with contaminated chlorhexidine 2% scrub in five domestic cats. J Vet Diagn Invest. 2018 ; 30:763–9.

46. Banfield Pet Hospital. Data shows increase in care for pets in 2020 despite pandemic. Available at: https://www.prnewswire.com/news-releases/banfield-pet-hospital-data-shows-increase-in-preventive-care-for-pets-in-2020-despite-pandemic-301204809.html. Accessed 4 February 2025.

47. Cooper LN, Beauchamp AM, Ingle TA, et al. Socioeconomic disparities and the prevalence of antimicrobial resistance. Clin Infect Dis. 2024 ; 79:1346–1353.

48. American Veterinary Medical Association. Antech and Idexx tout laboratory services, diagnostic innovations. Available at: https://www.avma.org/news/antech-and-idexx-tout-laboratory-services-diagnostic-innovations. Assessed 25 February 2025.

49. Burbick CR, Alexander TL, Wolking R, et al. Non-carbapenemase producing carbapenem-resistant *Klebsiella pneumoniae* isolated from the urinary tract of a dog. Can Vet J. 2022; 63:740–4.

50. Carter CN, Smith JL. A proposal to leverage high-quality veterinary diagnostic laboratory large data streams for animal health, public health, and One Health. J Vet Diagn Invest. 2021 ; 33:399–409.

