## Supplementary Table 1 for "Carbapenem-resistant Organisms in Companion Animals in New York City, 2019–2022"

**Supplementary Table 1**: Cleaning and categorization of source and site variables to create a single composite site variable from isolates identified by culture and antimicrobial susceptibility testing by animal care facilities in New York City from a single commercial diagnostic laboratory, 2019–2022.

**Composite Site Variable**

**Non-sterile sites**

| New variable | Original source variables |
|---|---|
| Abscess | Abscess |
| Aural (Ears) | Right ear (or AD), left ear (or AS), bulla, ear, ear canal |
| Gastrointestinal | Anal gland, bile, cecum, colon, duodenum, fecal, gallbladder, bile, rectal, vent, vulva |
| Intravenous catheter (IVC) | IVC |
| Lower respiratory | Bronchoalveolar lavage (BAL), bronchial, fluid tracheal, lung, trachea, transtracheal wash |
| Ocular | Conjunctiva, cornea, eye, right eye (or OD), left eye (or OS) |
| Oral | Oral, oral cavity, gingiva, mouth, tongue |
| Reproductive | Genital, penis, semen, testes, udder, uterus, vagina, vaginal discharge, vulva |
| Skin | Skin, bite, nails, nail bed, penile sheath, prepuce, hair |
| Soft tissue | Cyst, draining tract, fistula, mass, wound |
| Surgical site infection | Hardware, implant, incision site, surgery site, tibial plateau leveling osteotomy (TPLO) |
| Upper respiratory | Nasal, nasal discharge, nostril, oropharyngeal, pharynx, sinus, throat, respiratory |
| Urinary | Bladder stones, stones, bladder wall, catheterized, cystocentesis, free catch, ureter, urethra, urine |

**Sterile Sites**

| New variable | Original source variables |
|---|---|
| Blood | Blood |
| Bone | Bone |
| Cerebrospinal fluid (CSF) | CSF |
| Internal body site | Liver, kidney, omentum, lymph node, prostate, spleen |
| Joint | Joint |
| Pericardial & pericardial fluid | Fluid pericardial, pericardial |
| Peritoneal & peritoneal fluid | Abdominal fluid, ascites, fluid peritoneal, peritoneal |
| Pleural & pleural fluid | Fluid chest, fluid pleural, pleural, pyothorax |

**Other and Unknown**

| New variable | Original source variables |
|---|---|
| Other | Abdominal, aerobic swab, aspirate, biopsy, carpus carpal, elbow, leg, milk, other, pus, secretion, foot, neck, stifle, tail, thoracic, thorax, tissue, ulcer |
| Unknown | Unknown, missing data, source not indicated |
