## Supplementary Table 2 for "Carbapenem-resistant Organisms in Companion Animals in New York City, 2019–2022"

**Supplementary Table 2**: Distribution of gram-negative carbapenem-resistant organisms (CRO) and total gram-negative isolates identified by culture and antimicrobial susceptibility testing by animal care facilities in New York City from a single commercial diagnostic laboratory, by genus and species per patient species, 2019–2022.

| | Canine | | Feline | | Total | |
|---|---|---|---|---|---|---|
| | CRO n (%) | Gram-negative isolates n | CRO n (%) | Gram-negative isolates n | CRO n (%) | Gram-negative isolates n |
| Total Number of Bacterial Isolates | 190 (1.6) | 12,111 | 66 (1.6) | 4,004 | 256 (1.6) | 16,115 |
| Organism | | | | | | |
| *Bergeyella zoohelcum* | 0 | 2 | 1 (8.3) | 12 | 1 (7.1) | 14 |
| *Brevundimonas* spp. | 1 (9.1) | 11 | 0 | 3 | 1 (7.1) | 14 |
| *Brevundimonas diminuta* | 1 (100.0) | 1 | 0 | 0 | 1 (100.0) | 1 |
| *Brevundimonas vesicularis* | 0 | 3 | 0 | 0 | 0 | 3 |
| *Brevundimonas* spp. | 0 | 7 | 0 | 3 | 0 | 10 |
| *Burkholderia* spp. | 25 (55.6) | 45 | 12 (44.4) | 27 | 37 (51.4) | 72 |
| *Burkholderia cenocepacia* | 5 (100.0) | 5 | 7 (100.0) | 7 | 12 (100.0) | 12 |
| *Burkholderia cepacia* | 14 (60.9) | 23 | 3 (30.0) | 10 | 17 (51.5) | 33 |
| *Burkholderia* spp. | 6 (35.3) | 17 | 2 (20.0) | 10 | 8 (29.6) | 27 |
| *Chryseobacterium* spp. | 6 (60.0) | 10 | 5 (62.5) | 8 | 11 (61.1) | 18 |
| *Chryseobacterium gleum* | 2 (100.0) | 2 | 0 | 0 | 2 (100.0) | 2 |
| *Chryseobacterium indologenes* | 3 (100.0) | 3 | 3 (75.0) | 4 | 6 (85.7) | 7 |
| *Chryseobacterium* spp. | 1 (20.0) | 5 | 2 (50.0) | 4 | 3 (33.3) | 9 |
| *Citrobacter* spp. | 1 (1.6) | 62 | 0 | 22 | 1 (1.2) | 84 |
| *Citrobacter amalonaticus* | 0 | 2 | 0 | 2 | 0 | 4 |
| *Citrobacter braakii* | 0 | 5 | 0 | 0 | 0 | 5 |
| *Citrobacter farmeri* | 0 | 1 | 0 | 1 | 0 | 2 |
| *Citrobacter freundii* | 1 (3.8) | 26 | 0 | 13 | 1 (2.6) | 39 |
| *Citrobacter koseri* | 0 | 28 | 0 | 6 | 0 | 34 |
| *Elizabethkingia* spp. | 6 (85.7) | 7 | 2 (100.0) | 2 | 8 (88.9) | 9 |
| *Elizabethkingia meningoseptica* | 3 (75.0) | 4 | 1 (100.0) | 1 | 4 (80.0) | 5 |
| *Elizabethkingia* spp. | 3 (100.0) | 3 | 1 (100.0) | 1 | 4 (100.0) | 4 |
| *Empedobacter* spp. | 2 (100.0) | 2 | 0 | 0 | 2 (100.0) | 2 |
| *Enterobacter* spp. | 20 (7.2) | 278 | 8 (7.8) | 102 | 28 (7.4) | 380 |

| | | | | | | |
|---|---|---|---|---|---|---|
| *Enterobacter aerogenes* | 0 | 5 | 0 | 0 | 0 | 5 |
| *Enterobacter amnigenus* | 0 | 1 | 0 | 0 | 0 | 1 |
| *Enterobacter asburiae* | 0 | 0 | 0 | 1 | 0 | 1 |
| *Enterobacter cloacae* | 18 (11.4) | 158 | 7 (11.1) | 63 | 25 (11.3) | 221 |
| *Enterobacter hormaechei* | 0 | 1 | 0 | 0 | 0 | 1 |
| *Enterobacter* spp. | 2 (1.8) | 113 | 1 (2.6) | 38 | 3 (2.0) | 151 |
| *Escherichia* spp. | 28 (0.5) | 5759 | 12 (0.5) | 2513 | 40 (0.5) | 8272 |
| *Escherichia coli* | 28 (0.5) | 5736 | 12 (0.5) | 2509 | 40 (0.5) | 8245 |
| *Escherichia fergusonii* | 0 | 1 | 0 | 1 | 0 | 2 |
| *Escherichia hermannii* | 0 | 5 | 0 | 1 | 0 | 6 |
| *Escherichia vulneris* | 0 | 17 | 0 | 2 | 0 | 19 |
| *Klebsiella* spp. | 54 (6.7) | 804 | 15 (8.9) | 169 | 69 (7.1) | 973 |
| *Klebsiella aerogenes* | 0 | 2 | 0 | 0 | 0 | 2 |
| *Klebsiella oxytoca* | 0 | 14 | 0 | 2 | 0 | 16 |
| *Klebsiella pneumoniae* | 16 (9.0) | 178 | 5 (11.6) | 43 | 21 (9.5) | 221 |
| *Klebsiella pneumoniae* ssp *ozaenae* | 0 | 4 | 0 | 0 | 0 | 4 |
| *Klebsiella pneumoniae* ssp *pneumoniae* | 2 (13.3) | 15 | 0 | 7 | 2 (9.1) | 22 |
| *Klebsiella* spp. | 36 (6.1) | 591 | 10 (8.5) | 117 | 46 (6.5) | 708 |
| *Pantoea spp.* | 1 (0.7) | 136 | 0 | 22 | 1 (0.6) | 158 |
| *Pantoea agglomerans* | 0 | 5 | 0 | 0 | 0 | 5 |
| *Pantoea* spp. | 1 (0.8) | 131 | 0 | 22 | 1 (0.7) | 153 |
| *Pseudomonas* spp. | 42 (3.7) | 1143 | 10 (3.6) | 274 | 52 (3.7) | 1417 |
| *Pseudomonas aeruginosa* | 42 (4.3) | 986 | 10 (4.5) | 223 | 52 (4.3) | 1209 |
| *Pseudomonas alcaligenes* | 0 | 1 | 0 | 0 | 0 | 1 |
| *Pseudomonas fluorescens* | 0 | 2 | 0 | 0 | 0 | 2 |
| *Pseudomonas luteola* | 0 | 2 | 0 | 3 | 0 | 5 |
| *Pseudomonas putida* | 0 | 19 | 0 | 5 | 0 | 24 |
| *Pseudomonas stutzeri* | 0 | 5 | 0 | 0 | 0 | 5 |
| *Pseudomonas* spp. | 0 | 128 | 0 | 43 | 0 | 171 |
| *Ralstonia* spp. | 0 | 7 | 1 (50.0) | 2 | 1 (11.1) | 9 |
| *Ralstonia insidiosa* | 0 | 1 | 1 (100.0) | 1 | 1 (50.0) | 2 |
| *Ralstonia pickettii* | 0 | 6 | 0 | 1 | 0 | 7 |
| *Sphingobacterium* spp. | 3 (75.0) | 4 | 0 | 0 | 3 (75.0) | 4 |
| *Sphingobacterium multivorum* | 3 (100.0) | 3 | 0 | 0 | 3 (100.0) | 3 |
| *Sphingobacterium* spp. | 0 | 1 | 0 | 0 | 0 | 1 |
| *Yersinia* spp. | 1 (33.3) | 3 | 0 | 0 | 1 (33.3) | 3 |
| *Yersinia enterocolitica* | 1 (50.0) | 2 | 0 | 0 | 1 (50.0) | 2 |
| *Yersinia pseudotuberculosis* | 0 | 1 | 0 | 0 | 0 | 1 |
