## Supplementary Table 3 for "Carbapenem-resistant Organisms in Companion Animals in New York City, 2019–2022"

**Supplementary Table 3:** Count and percentage of isolates of carbapenem-resistant organisms (CRO) identified by culture and antimicrobial susceptibility testing by animal care facilities in New York City from a single commercial diagnostic laboratory, by specimen site[a] and genus, 2019–2022.

| | | Non-sterile sites | | | | | | | | Normally sterile sites[c] n (%) | Other[d] n (%) | Unknown n (%) |
|---|---|---|---|---|---|---|---|---|---|---|---|---|
| Total isolates | | Abscess n (%) | Ears n (%) | Lower Respiratory n (%) | Upper Respiratory n (%) | Skin & Soft tissue n (%) | Surgical site infections n (%) | Urinary n (%) | Other Non-Sterile Sites[b] n (%) | | | |
| Organisms | | | | | | | | | | | | |
| *Bergeyella zoohelcum* | CRO (n=1) | 0 | 0 | 0 | 1 (100.0) | 0 | 0 | 0 | 0 | 0 | 0 | 0 |
| | Total (n=14) | 0 | 0 | 7 (50.0) | 7 (50.0) | 0 | 0 | 0 | 0 | 0 | 0 | 0 |
| *Brevundimonas diminuta* | CRO (n=1) | 0 | 0 | 0 | 0 | 0 | 1 (100.0) | 0 | 0 | 0 | 0 | 0 |
| | Total (n=14) | 0 | 1 (7.1) | 3 (21.4) | 0 | 4 (28.6) | 1 (7.1) | 2 (14.3) | 0 | 0 | 1 (7.1) | 2 (14.3) |
| *Burkholderia* spp. | CRO (n=37) | 4 (10.8) | 12 (32.4) | 3 (8.1) | 0 | 12 (32.4) | 0 | 4 (10.8) | 1 (2.7) | 0 | 0 | 1 (2.7) |
| | Total (n=72) | 4 (5.6) | 17 (23.6) | 6 (8.3) | 4 (5.6) | 23 (31.9) | 0 | 12 (16.7) | 2 (2.8) | 1 (1.4) | 2 (2.8) | 1 (1.4) |
| *Chryseobacterium* spp. | CRO (n=11) | 1 (9.1) | 1 (9.1) | 2 (18.2) | 2 (18.2) | 3 (27.3) | 0 | 0 | 1 (9.1) | 0 | 1 (9.1) | 0 |
| | Total (n=18) | 1 (5.6) | 3 (16.7) | 3 (16.7) | 3 (16.7) | 6 (33.3) | 0 | 0 | 1 (5.6) | 0 | 1 (5.6) | 0 |
| *Citrobacter freundii* | CRO (n=1) | 0 | 0 | 1 (100.0) | 0 | 0 | 0 | 0 | 0 | 0 | 0 | 0 |
| | Total (n=84) | 2 (2.4) | 9 (10.7) | 4 (4.8) | 5 (6.0) | 13 (15.5) | 1 (1.2) | 44 (52.4) | 5 (5.6) | 0 | 1 (1.2) | 0 |
| *Elizabethkingia* spp. | CRO (n=8) | 1 (12.5) | 0 | 1 (12.5) | 1 (12.5) | 3 (37.5) | 0 | 1 (12.5) | 1 (12.5) | 0 | 0 | 0 |
| | Total (n=9) | 1 (11.1) | 0 | 1 (11.1) | 1 (11.1) | 3 (33.3) | 0 | 1 (11.1) | 1 (11.1) | 1 (11.1) | 0 | 0 |
| *Empedobacter* spp. | CRO (n=2) | 0 | 0 | 0 | 0 | 2 (100.0) | 0 | 0 | 0 | 0 | 0 | 0 |
| | Total (n=2) | 0 | 0 | 0 | 0 | 2 (100.0) | 0 | 0 | 0 | 0 | 0 | 0 |
| *Enterobacter* spp. | CRO (n=28) | 2 (7.1) | 0 (0.0) | 4 (14.3) | 1 (3.6) | 4 (14.3) | 2 (7.1) | 6 (21.4) | 2 (7.1) | 3 (10.7) | 4 (14.3) | 0 |

| | | | | | | | | | | | | |
|---|---|---|---|---|---|---|---|---|---|---|---|---|
| | Total (n=380) | 21 (5.5) | 17 (4.5) | 22 (5.8) | 15 (4.0) | 91 (24.0) | 7 (1.8) | 165 (43.4) | 17 (4.5) | 14 (3.7) | 9 (2.4) | 2 (0.5) |
| *Escherichia coli* | CRO (n=40) | 1 (2.5) | 2 (5.0) | 4 (10.0) | 1 (2.5) | 4 (10.0) | 1 (2.5) | 17 (42.5) | 4 (10.0) | 5 (12.5) | 1 (2.5) | 0 |
| | Total (n=8272) | 127 (1.5) | 357 (4.3) | 87 (1.1) | 78 (0.9) | 409 (4.9) | 36 (0.4) | 6526 (78.9) | 375 (4.5) | 93 (1.1) | 72 (0.9) | 112 (1.4) |
| *Klebsiella* spp. | CRO (n=69) | 3 (4.4) | 0 (0.0) | 18 (26.1) | 3 (4.4) | 7 (10.1) | 7 (10.1) | 20 (29.0) | 3 (4.4) | 1 (1.5) | 6 (8.7) | 1 (1.5) |
| | Total (n=973) | 32 (3.3) | 54 (5.6) | 53 (5.5) | 21 (2.3) | 113 (11.6) | 22 (2.3) | 599 (61.6) | 39 (4.0) | 11 (1.1) | 18 (1.9) | 10 (1.0) |
| *Pantoea* spp. | CRO (n=1) | 0 | 0 | 0 | 0 | 1 (100.0) | 0 | 0 | 0 | 0 | 0 | 0 |
| | Total (n=158) | 10 (6.3) | 21 (13.3) | 1 (0.6) | 4 (2.5) | 71 (44.9) | 1 (0.6) | 39 (24.7) | 4 (2.5) | 0 (0.0) | 5 (3.2) | 2 (1.3) |
| *Pseudomonas aeruginosa* | CRO (n=52) | 1 (1.9) | 28 (53.9) | 1 (1.9) | 9 (17.3) | 7 (13.5) | 1 (1.9) | 4 (7.7) | 0 | 0 | 1 (1.9) | 0 |
| | Total (n=1417) | 52 (3.7) | 573 (40.4) | 42 (3.0) | 123 (8.7) | 318 (22.4) | 19 (1.3) | 196 (13.8) | 52 (3.7) | 12 (0.9) | 16 (1.1) | 14 (1.0) |
| *Ralstonia insidiosa* | CRO (n=1) | 0 | 0 | 0 | 0 | 0 | 0 | 0 | 1 (100.0) | 0 | 0 | 0 |
| | Total (n=9) | 0 | 0) | 7 (77.8) | 0 | 0 | 0 | 0 | 1 (11.1) | 0 | 1 (11.1) | 0 |
| *Sphingobacterium multivorum* | CRO (n=3) | 0 | 1 (33.3) | 0 | 0 | 1 (33.3) | 1 (33.3) | 0 | 0 | 0 | 0 | 0 |
| | Total (n=4) | 0 | 1 (25.0) | 0 | 0 | 2 (5.0) | 1 (25.0) | 0 | 0 | 0 | 0 | 0 |
| *Yersinia enterocolitica* | CRO (n=1) | 0 | 0 | 0 | 0 | 0 | 0 | 0 | 0 | 0 | 0 | 1 (100.0) |
| | Total (n=3) | 1 (33.3) | 0 | 0 | 0 | 1 (33.3) | 0 | 0 | 0 | 0 | 0 | 1 (33.3) |
| Total | CRO (n=256) | 13 (5.1) | 44 (17.2) | 34 (13.3) | 18 (7.0) | 44 (17.2) | 13 (5.1) | 52 (20.3) | 13 (5.1) | 9 (3.5) | 13 (5.1) | 3 (1.2) |
| | Total (n=16,115) | 470 (2.9) | 1770 (11.0) | 404 (2.5) | 469 (2.9) | 1914 (11.9) | 127 (0.8) | 9734 (60.4) | 676 (4.2) | 178 (1.1) | 176 (1.1) | 197 (1.2) |

[a]Composite site variable was used for these data.
[b] Other non-sterile sites included abscesses, gastrointestinal, ocular, oral, and reproductive sites.
[c] Normally sterile sites included blood, cerebrospinal fluid, and samples taken from peritoneal, pericardial, and pleural sites, other internal body sites, joints, and bone.
[d] "Other" included sites that could not be clearly categorized into the listed sites.
